# Cognitive Impairment Among People with Epilepsy in Peru

**DOI:** 10.64898/2026.08.28.26361672

**Authors:** Samantha E. Allen, Christopher Phillips, Melissa T. Wardle, Luz M. Moyano, Javier A. Bustos, Luis L. Rojas, Narcisa R. Otero, Lucia Bolivar-Herrada, Seth E. O’Neal, Héctor H. García, the Cysticercosis Working Group in Peru (CWGP)

## Abstract

**Objective:** Cognitive impairment is a common comorbidity among people with epilepsy (PWE) and is associated with disability and reduced quality of life. We characterized the burden of cognitive impairment and identified factors associated with cognitive performance in a large, population-based cohort of PWE living in Northern Peru, a region highly endemic for *Taenia solium* where neurocysticercosis (NCC) is a common cause of acquired epilepsy.

**Methods:** PWE enrolled in a population-based cohort in Northern Peru between 2007 and 2020 completed the Mini-Mental State Examination (MMSE) at enrollment. Cognitive impairment was defined as an MMSE score <24. Demographic and clinical data, including epilepsy characteristics and NCC status, were collected. Negative binomial regression was used to identify factors associated with the number of MMSE errors.

**Results:** Among 764 participants, the mean MMSE score was 26.4 (SD 4.2), and 16.4% met criteria for cognitive impairment. Memory and attention were the most affected domains. In multivariable analysis, older age and lower educational attainment were independently associated with poorer cognitive performance.

**Conclusion:** In this large, community-based cohort from Northern Peru, approximately 1 in 6 PWE had abnormal global cognition on the MMSE, with memory and attention most affected. These findings underscore the importance of incorporating cognitive evaluation and management into comprehensive epilepsy care, particularly in resource-limited settings where cognitive morbidity may be underrecognized. Given the potential for cognitive difficulties to compound disability and adversely affect quality of life, identifying and addressing cognitive morbidity may be especially important in populations already facing substantial barriers to epilepsy care.

## 1. Introduction

Cognitive deficits are believed to afflict up to 80% of people with epilepsy (PWE) and are increasingly recognized as a major contributor to disability and reduced quality of life in this population.^1,2^ Although recurrent seizures may exacerbate cognitive dysfunction, it is now recognized that seizures and cognitive impairment often arise from a shared underlying neuropathology rather than a simple cause-and-effect relationship.^2,3^ Additional contributors include genetic susceptibility, psychiatric and medical comorbidities, psychosocial and socioeconomic factors, and treatment-related adverse effects.^4–6^ This complex interplay of factors makes cognitive impairment both difficult to predict and challenging to treat. Consequently, epilepsy care has evolved beyond seizure control alone to encompass cognitive and psychological outcomes as essential components of comprehensive management.^7,8^ Indeed, for many PWE, cognitive impairment may have a greater impact on daily functioning than seizures themselves. Accurately characterizing the burden and determinants of cognitive impairment is therefore essential to informing comprehensive epilepsy care and identifying opportunities for intervention.

The link between cognitive dysfunction and quality of life, while incompletely characterized, has been consistently demonstrated. PWE often experience reduced social participation due to the combined effects of chronic illness and societal stigma.^9,10^ Cognitive impairment may further compound these challenges by affecting memory, attention, processing speed, and executive functioning, making it more difficult to communicate effectively, maintain interpersonal relationships, and engage in work, school, and community activities.^4–6^ Consequently, cognitive dysfunction may be among the most important determinants of quality of life in PWE, independent of seizure burden.^1^

While cognitive dysfunction is an important consideration for PWE worldwide, those living in low- and middle-income countries (LMICs) face additional, unique challenges that may increase the risk of cognitive impairment.^11,12^ In these resource-limited settings, access to newer antiseizure medications (ASMs) is often constrained, leading to greater reliance on older agents with less favorable profiles of cognitive adverse effects. In particular, barbiturates and benzodiazepines, medications rarely used as first-line therapies in high-income settings because of their well-recognized cognitive side effects, remain among the most widely available ASMs in many LMICs.^13,14^ Beyond differences in ASM availability, resource constraints may also limit timely diagnosis, access to comprehensive epilepsy care, cognitive assessment, and long-term follow-up, potentially compounding the burden of cognitive dysfunction.^15^

In addition to healthcare resource limitations, many LMICs also bear a disproportionate burden of acquired epilepsies from varied etiologies including traumatic brain injuries and infections that are well known to independently contribute to cognitive dysfunction.^16^ Among these, neurocysticercosis (NCC), caused by infection with the larval stage of *Taenia solium*, is one of the leading causes of acquired epilepsy in endemic regions.^17^ The clinical manifestations of NCC are highly heterogeneous and are influenced by parasite-related factors, including the number, location, and stage of cysts, as well as the host immune response.^18^ Although seizures are the most well-recognized manifestation of NCC, the disease is a multisystem neurologic disorder with a broad spectrum of clinical manifestations. ^19^ Cognitive impairment is a common and often underrecognized source of morbidity, ^20–23^ frequently accompanied by psychiatric comorbidities such as mood and anxiety disorders that may further exacerbate cognitive dysfunction and diminish quality of life. ^23–27^

Despite this growing recognition of cognitive impairment in epilepsy, most existing data are derived from high-income countries, and the burden and determinants of cognitive dysfunction among PWE in LMIC remain poorly characterized. To address this gap, we conducted a cross-sectional study nested within a longitudinal epilepsy cohort in a *T. solium*-endemic region of northern Peru to characterize the prevalence, pattern, and determinants of cognitive impairment in this population.

## 2. Methods

### 2.1 Study design and setting

Participants are drawn from a prospective cohort established in Tumbes, Peru, a highly endemic area for *T. solium*, designed to follow PWE. The motivation behind forming this cohort was to better understand this population and elucidate outcomes for PWE in this context. The cohort formation and baseline demographics have been published previously.^28^ In brief, study participants were drawn from 107 villages within the state of Tumbes, Peru, as well as from the neighboring state of Piura. Participants were screened through home visits for symptoms related to epilepsy using a validated questionnaire.^29^ Those who screened positive were referred to the Center for Global Health in Tumbes, Peru, for in-person clinician evaluations. Self-referrals were also accepted. Screening and cohort enrollment occurred from 2007 to 2020 through the Center for Global Health in Tumbes, Peru.

### 2.2 Study participants and evaluation

Inclusion criteria included the following: (1) greater than 2 years of age; (2) a diagnosis of epilepsy, defined as two or more unprovoked seizures separated by over 24 hours; and (3) residence in a *T. solium* endemic region. A total of 1975 individuals completed baseline evaluations, including a clinical questionnaire and an in-person clinical evaluation, as well as diagnostics such as CT neuroimaging, serologies, and EEG. The present analysis includes a subset of the population that completed the Mini-Mental Status Exam (MMSE).

### 2.3 Cognitive evaluation

The MMSE was administered to participants who consented to the additional battery of complementary tests. The MMSE is an 11-question validated clinical cognitive screening test, widely used throughout Latin America,^30^ covering 5 domains including orientation, language/praxis, attention, immediate recall/registration, and delayed recall.^31^ The test usually takes between 10 and 15 minutes to complete and is scored from 0 to 30, with lower scores correlating with increased cognitive impairment. A cut-off composite score of less than 24 out of 30 was used to indicate cognitive impairment. For the present study, the test was administered in-person and scored by specially trained research personnel during the baseline evaluation cohort visit (LBH, CAL, BIC, and LHM). Orientation was calculated as the sum of temporal and spatial orientation items (range 0–10). Registration was assessed using the immediate recall component (range 0–3). Attention was assessed using the attention/calculation component (range 0–5). Memory was assessed using delayed recall (range 0–3). Language was calculated as the sum of naming, repetition, comprehension (three-step command), reading, writing, and figure-copying items (range 0–9) (Table S1).

### 2.4 NCC diagnosis

A diagnosis of NCC was made using the principles of the Del Brutto criteria. ^32^ In brief, by virtue of living in this region and having a diagnosis of epilepsy, all participants already met two of the minor clinical/exposure Del Brutto criteria. To have at least a “probable diagnosis” of NCC, participants also needed neuroimaging demonstrating active or chronic NCC (cystic lesions with or without a discernible scolex, enhancing lesions, multilobulated cystic lesions in the subarachnoid space, and/or typical parenchymal calcifications). For this analysis, participants’ NCC status was dichotomized based on imaging findings: those with neuroimaging evidence of active or chronic NCC were considered to have NCC, and those without imaging biomarkers of NCC were classified as negative for NCC.

### 2.5 Data analysis

Descriptive statistics were used to characterize the study population overall and according to MMSE performance, categorized as normal (MMSE ≥24) or impaired (MMSE <24). Demographic, clinical, and epilepsy-related characteristics were compared between groups using chi-square or Fisher’s exact tests for categorical variables and independent-samples t-tests for continuous variables. Because individual MMSE cognitive domains have different maximum possible scores, scores were expressed as percentages of the maximum possible score for each domain to facilitate comparison across domains, and are shown with 95% confidence intervals.

Negative binomial regression was used as the primary multivariable analysis to evaluate factors associated with the number of MMSE errors. The distribution of MMSE errors was highly right-skewed and demonstrated substantial overdispersion relative to the mean; therefore, negative binomial regression was selected over Poisson regression. Results are presented as rate ratios (IRRs) with 95% confidence intervals, with IRRs representing the relative difference in the expected number of MMSE errors associated with each predictor. The multivariable model was adjusted for age, educational attainment, duration of epilepsy, and NCC diagnosis. Unadjusted models included all participants with available data for the respective predictor, whereas the adjusted model was restricted to participants with complete data for all covariates. Candidate variables were selected based on associations in univariate analyses (*p* < 0.10) and established associations reported in the literature. Variables with more than 20% missing data were excluded from regression analyses. NCC was included in the model based on its established association with cognitive impairment. ^20,22,33,34^ Because current age, age at epilepsy onset, and epilepsy duration are mathematically related, these variables were not included simultaneously in the primary model. Alternative multivariable model specifications were evaluated to assess the independent associations of age at epilepsy onset and epilepsy duration with cognitive performance. Separate models included either age at epilepsy onset or epilepsy duration, along with current age and other covariates. An α-level of 0.05 was used to determine statistical significance. All analyses were conducted using Stata version 18 (StataCorp LLC, College Station, TX).

## 3. Results

A total of 764 participants (38.7%) completed the MMSE among the 1,975 individuals enrolled in the cohort. The mean age of participants who completed the MMSE was 30.3 years (SD 15.0), and female participants slightly outnumbered male participants overall and across MMSE performance groups. The mean MMSE score was 26.3 (SD 4.2), with cognitive impairment (MMSE < 24) identified in 16.4% of participants (n=132). Participants with cognitive impairment were significantly older than those without impairment (mean age 36.7 years [SD 17.8] vs. 29.0 years [SD 14.1], *p* < 0.001). The proportion of female participants did not differ significantly by MMSE performance group (*p* > 0.05) (Table 1).

**Table 1.**
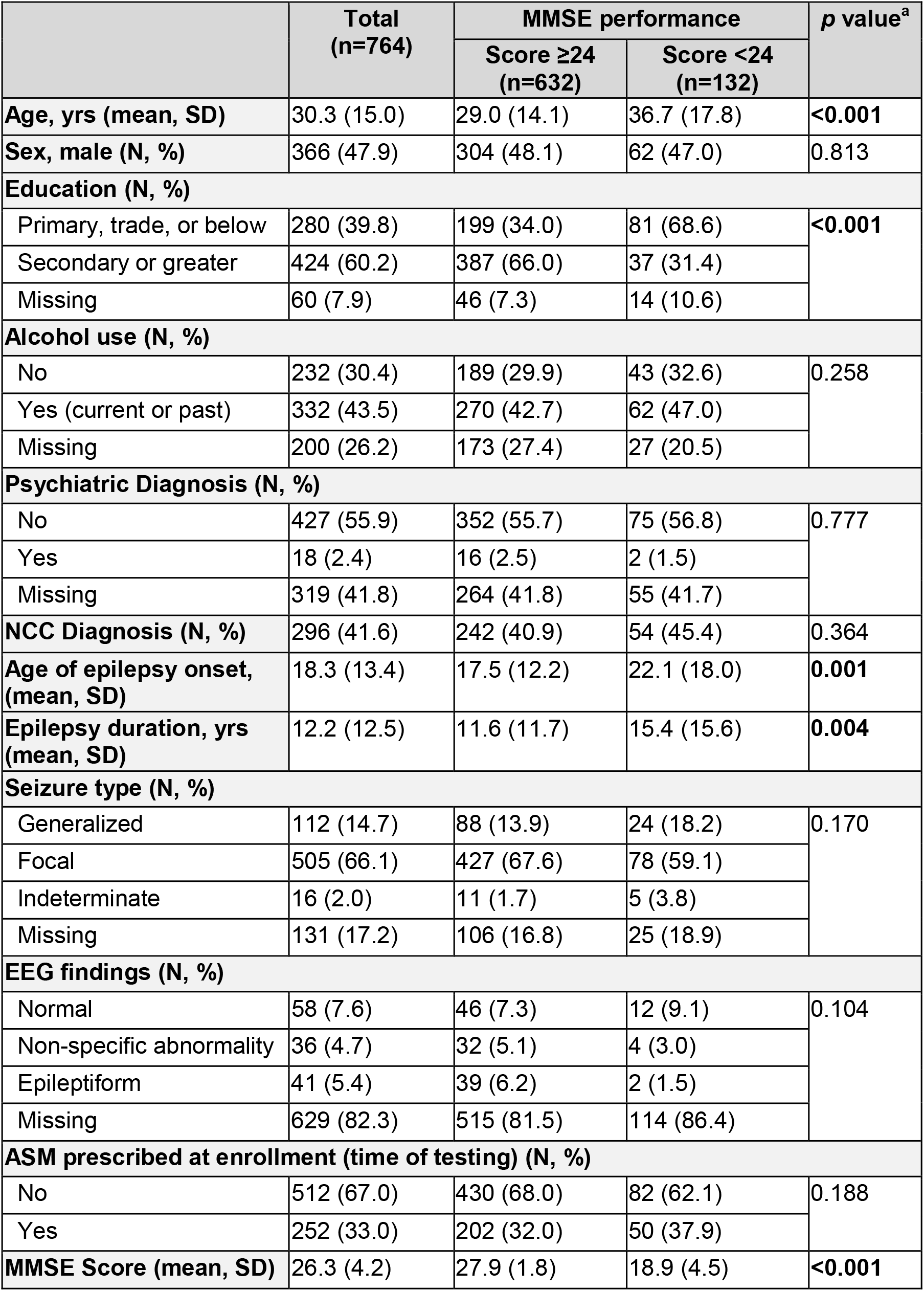
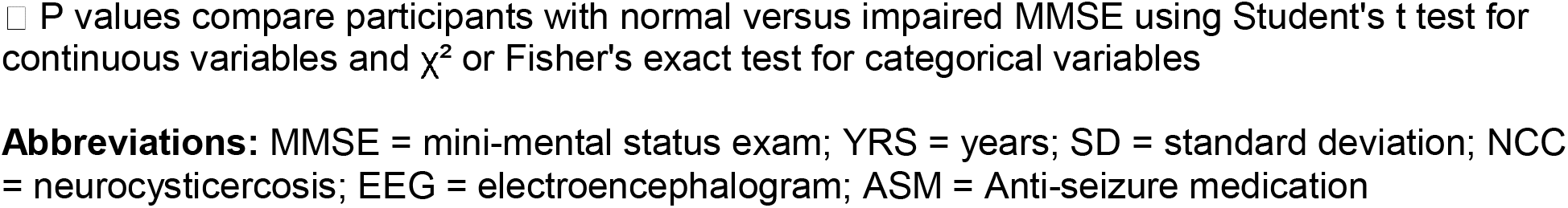
Population Baseline Characteristics.

|  | Total<br>(n=764) | MMSE performance |  | p value<sup>a</sup> |
| --- | --- | --- | --- | --- |
|  |  | Score ≥24<br>(n=632) | Score <24<br>(n=132) |  |
| Age, yrs (mean, SD) | 30.3 (15.0) | 29.0 (14.1) | 36.7 (17.8) | <0.001 |
| Sex, male (N, %) | 366 (47.9) | 304 (48.1) | 62 (47.0) | 0.813 |
| Education (N, %) |  |  |  |  |
| Primary, trade, or below | 280 (39.8) | 199 (34.0) | 81 (68.6) | <0.001 |
| Secondary or greater | 424 (60.2) | 387 (66.0) | 37 (31.4) |  |
| Missing | 60 (7.9) | 46 (7.3) | 14 (10.6) |  |
| Alcohol use (N, %) |  |  |  |  |
| No | 232 (30.4) | 189 (29.9) | 43 (32.6) | 0.258 |
| Yes (current or past) | 332 (43.5) | 270 (42.7) | 62 (47.0) |  |
| Missing | 200 (26.2) | 173 (27.4) | 27 (20.5) |  |
| Psychiatric Diagnosis (N, %) |  |  |  |  |
| No | 427 (55.9) | 352 (55.7) | 75 (56.8) | 0.777 |
| Yes | 18 (2.4) | 16 (2.5) | 2 (1.5) |  |
| Missing | 319 (41.8) | 264 (41.8) | 55 (41.7) |  |
| NCC Diagnosis (N, %) | 296 (41.6) | 242 (40.9) | 54 (45.4) | 0.364 |
| Age of epilepsy onset, (mean, SD) | 18.3 (13.4) | 17.5 (12.2) | 22.1 (18.0) | 0.001 |
| Epilepsy duration, yrs (mean, SD) | 12.2 (12.5) | 11.6 (11.7) | 15.4 (15.6) | 0.004 |
| Seizure type (N, %) |  |  |  |  |
| Generalized | 112 (14.7) | 88 (13.9) | 24 (18.2) | 0.170 |
| Focal | 505 (66.1) | 427 (67.6) | 78 (59.1) |  |
| Indeterminate | 16 (2.0) | 11 (1.7) | 5 (3.8) |  |
| Missing | 131 (17.2) | 106 (16.8) | 25 (18.9) |  |
| EEG findings (N, %) |  |  |  |  |
| Normal | 58 (7.6) | 46 (7.3) | 12 (9.1) | 0.104 |
| Non-specific abnormality | 36 (4.7) | 32 (5.1) | 4 (3.0) |  |
| Epileptiform | 41 (5.4) | 39 (6.2) | 2 (1.5) |  |
| Missing | 629 (82.3) | 515 (81.5) | 114 (86.4) |  |
| ASM prescribed at enrollment (time of testing) (N, %) |  |  |  |  |
| No | 512 (67.0) | 430 (68.0) | 82 (62.1) | 0.188 |
| Yes | 252 (33.0) | 202 (32.0) | 50 (37.9) |  |
| MMSE Score (mean, SD) | 26.3 (4.2) | 27.9 (1.8) | 18.9 (4.5) | <0.001 |

□ P values compare participants with normal versus impaired MMSE using Student's t test for continuous variables and $\chi^2$ or Fisher's exact test for categorical variables
**Abbreviations:** MMSE = mini-mental status exam; YRS = years; SD = standard deviation; NCC = neurocysticercosis; EEG = electroencephalogram; ASM = Anti-seizure medication

Approximately 60% of participants had attained a secondary or higher level of education, although educational attainment was substantially lower among those with impaired cognition (31.4% vs. 66.0%, *p* < 0.001). Psychiatric diagnoses were relatively uncommon, reported in 2.4% of the overall population, while alcohol use was reported by more than 40% of participants. Both alcohol use and psychiatric diagnosis had more than 20% missing observations (Table 1).

Approximately two-thirds of participants (66%) had focal seizures. The mean age at epilepsy onset was 18.3 years (SD 13.4), with a mean epilepsy duration of 12.2 years (SD 12.5).

Compared with participants without cognitive impairment, those with impaired cognition had a later mean age at epilepsy onset (22.1 vs. 17.5 years, *p* = 0.001) and longer mean epilepsy duration (15.4 vs. 11.6 years, *p* = 0.004). No significant differences between groups were observed in alcohol use, history of psychiatric diagnosis, seizure type, EEG findings, or ASM use. EEG findings were available for only 17.7% of participants, limiting comparisons between groups. Notably, only 33% of participants had been prescribed an ASM prior to study enrollment (Table 1).

Neuroimaging showed NCC in 41.6% of participants, the vast majority of whom had calcified lesions. Mean MMSE scores did not differ between participants with and without NCC, either overall or across individual cognitive domains (Table 1).

Across the cohort, MMSE performance varied across cognitive domains, with the highest mean percentage scores observed for registration (97.9%, 95% CI 97.1–98.8) and orientation (91.7%, 95% CI 90.6–92.8). Lower performance was observed in language/praxis (89.1%, 95% CI 87.9– 90.3), attention (80.2%, 95% CI 77.9–82.6), and memory (77.3%, 95% CI 74.9–79.7), with memory and attention showing the lowest mean percentage scores (Figure 1).

**Figure 1.**
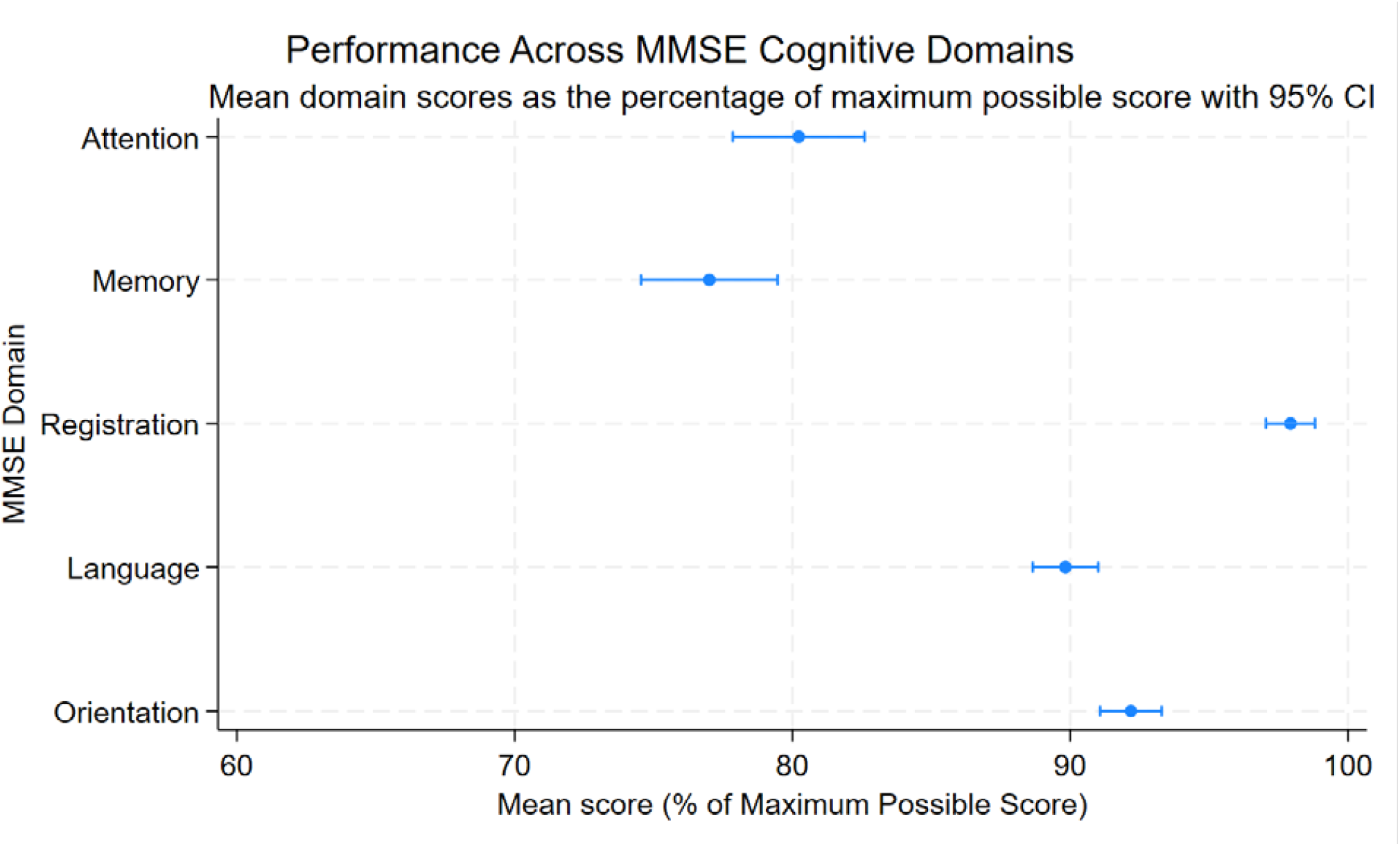
Distribution of Mini-Mental Status Exam (MMSE) scores by Domain

Variables considered for the multivariable models included age, duration of epilepsy, educational attainment, age at epilepsy onset, and NCC. Alcohol use and psychiatric diagnosis were not included in the primary multivariable models because both had more than 20% missing observations. Because current age, age at epilepsy onset, and epilepsy duration demonstrated substantial multicollinearity, separate models were examined that included either age at epilepsy onset or epilepsy duration.

Of the 764 participants who completed the MMSE, 567 had complete data for all covariates included in the primary multivariable model. In the adjusted negative binomial regression model, participants with primary education or less had a greater expected number of MMSE errors than those with secondary or greater education (IRR 1.87, 95% CI 1.57–2.22; *p* < 0.001), corresponding to an estimated 87% higher number of errors. Older age was modestly associated with a greater number of MMSE errors (IRR 1.007 per year, 95% CI 1.000–1.013; *p* = 0.049), whereas epilepsy duration and NCC were not independently associated with the number of MMSE errors (Table 2).

**Table 2.**
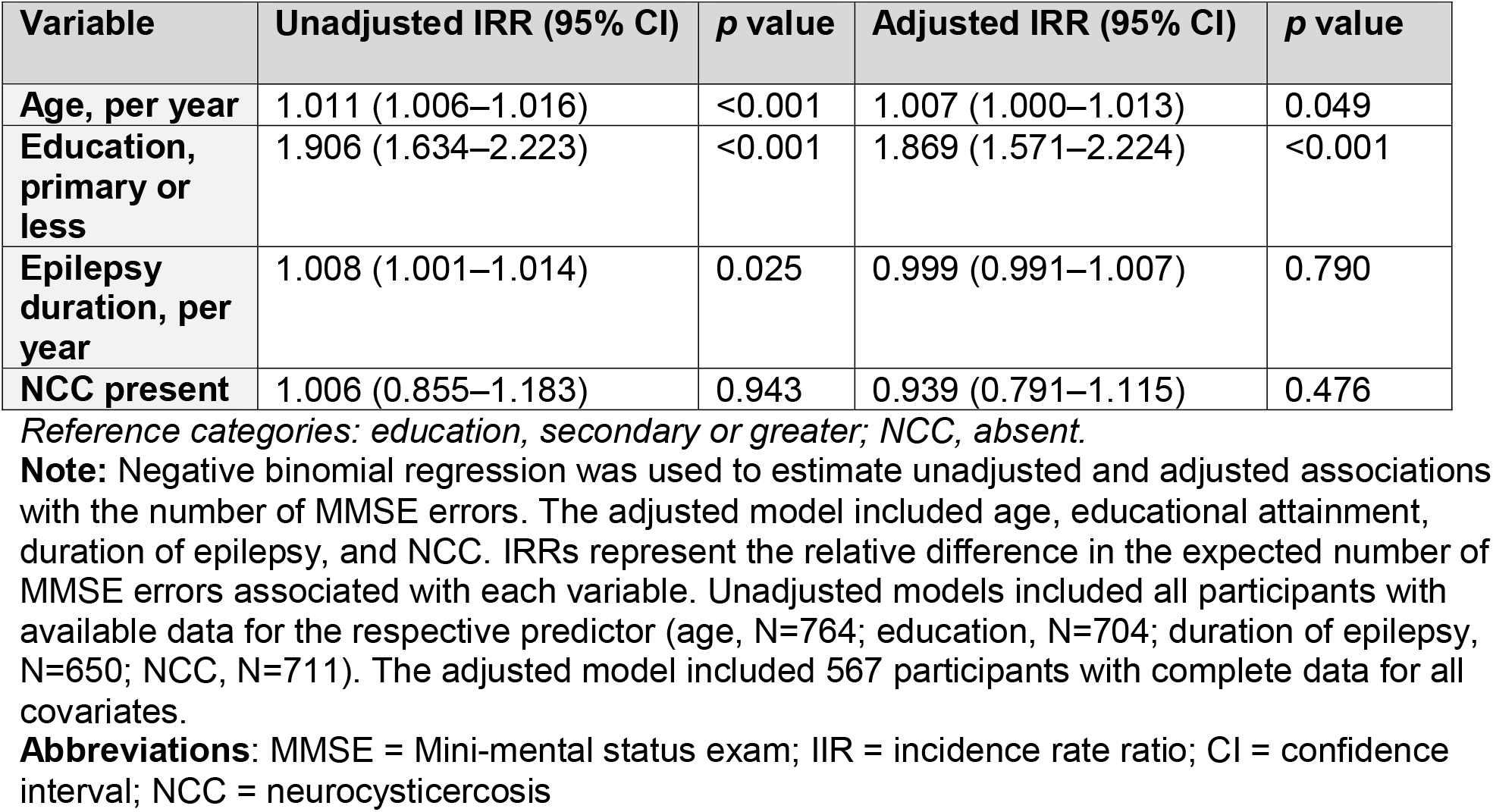
Factors associated with number of MMSE errors among people with epilepsy.

| Variable | Unadjusted IRR (95% CI) | p value | Adjusted IRR (95% CI) | p value |
| --- | --- | --- | --- | --- |
| Age, per year | 1.011 (1.006–1.016) | <0.001 | 1.007 (1.000–1.013) | 0.049 |
| Education, primary or less | 1.906 (1.634–2.223) | <0.001 | 1.869 (1.571–2.224) | <0.001 |
| Epilepsy duration, per year | 1.008 (1.001–1.014) | 0.025 | 0.999 (0.991–1.007) | 0.790 |
| NCC present | 1.006 (0.855–1.183) | 0.943 | 0.939 (0.791–1.115) | 0.476 |

*Reference categories: education, secondary or greater; NCC, absent.*
**Note:** Negative binomial regression was used to estimate unadjusted and adjusted associations with the number of MMSE errors. The adjusted model included age, educational attainment, duration of epilepsy, and NCC. IRRs represent the relative difference in the expected number of MMSE errors associated with each variable. Unadjusted models included all participants with available data for the respective predictor (age, N=764; education, N=704; duration of epilepsy, N=650; NCC, N=711). The adjusted model included 567 participants with complete data for all covariates.
**Abbreviations:** MMSE = Mini-mental status exam; IIR = incidence rate ratio; CI = confidence interval; NCC = neurocysticercosis

In an alternative multivariable model replacing epilepsy duration with age at epilepsy onset, age at epilepsy onset was not independently associated with the number of MMSE errors (IRR 1.001, 95% CI 0.993–1.008; *p* = 0.879). The association between lower educational attainment and a greater number of MMSE errors remained robust, whereas NCC was not associated with MMSE errors (data not shown).

## 4. Conclusions

In this cohort of PWE from northern Peru, cognitive impairment was common, with approximately 1 in 6 participants demonstrating abnormal global cognition on the MMSE, suggesting that cognitive impairment represents an important comorbidity among PWE in this setting. The most common domains of impairment were memory and attention, evaluated using delayed recall and serial 7s testing, respectively. Lower educational attainment was strongly associated with a greater number of MMSE errors; participants with primary education or less had an estimated 87% higher number of errors than those with secondary education or higher. Older age was more modestly associated with a greater number of MMSE errors. Although longer epilepsy duration and older age at epilepsy onset were associated with poorer cognitive performance in univariate analyses, neither remained independently associated with MMSE errors after adjustment.

The observed associations between MMSE performance and age, educational attainment, age at epilepsy onset, and epilepsy duration have been well described in the literature.^36,37^ Numerous studies have shown age-related decline in cognitive performance, both in the general population and among individuals with epilepsy.^38^ Recent work in another Peruvian cohort with calcified NCC similarly identified age as one of the strongest predictors of lower cognitive screening scores.^39^ Educational attainment corresponding to higher scores is also expected given the well-recognized influence of education on cognitive screening instruments, particularly the MMSE, which is sensitive to literacy and educational opportunity.^40^

No difference in performance was found between participants with and without NCC. Prior studies have demonstrated a spectrum of cognitive abnormalities in NCC, ranging from subtle deficits in attention and executive functioning to more severe cognitive impairment.^22,41,42^ Del Brutto and others have shown that calcified NCC may be associated with hippocampal atrophy and mesial temporal sclerosis,^43,44^ and Varghese et al. reported that even patients with inactive NCC lesions exhibited impaired performance on memory and attention tasks compared with controls.^20^ The literature is, however, heterogeneous, likely reflecting a range of differences in disease stage and severity, seizure control and associated co-morbidities, and the cognitive instruments used for assessment. Some studies have similarly found little or no difference in global cognitive screening measures between PWE with NCC and those without.^45–47^ Importantly, all participants in our study are PWE. Thus, our comparison assesses whether NCC is associated with additional differences in global cognitive performance among PWE, rather than relative to neurologically healthy controls. Because epilepsy itself may contribute to cognitive impairment, this may limit the ability to detect an independent effect of NCC.

The absence of an observed association between NCC and cognitive performance in our study should not be interpreted as evidence that NCC has no cognitive consequences. Rather, our findings suggest that among PWE, the presence of NCC alone may not be a primary determinant of cognitive performance. NCC nevertheless likely influences cognition indirectly, either through its contribution to epilepsy or through disease characteristics not captured in this study. Further, the MMSE is a relatively brief screening instrument and may not capture the subtle deficits in specific cognitive domains described in NCC.

The high treatment gap in this setting,^28^ with two-thirds not prescribed an ASM prior to enrollment, provides an important context for interpreting these findings. Limited access to treatment with a range of ASMs as well as other neurologic and psychiatric services may contribute to persistent seizures and undertreated comorbidities, potentially increasing the overall burden of cognitive difficulties in this population. In this context, addressing cognitive impairment may require a broader approach than cognitive assessment alone, including improved access to epilepsy specific treatments and identification and management of psychiatric comorbidities. Longitudinal studies will be needed to determine whether improving seizure control and access to mental health and comprehensive neurologic care leads to improved cognitive outcomes in this population.

Several limitations should be considered. First, the study relies on screening-level cognitive assessment using the MMSE rather than full neuropsychological batteries, which may under-detect subtle deficits and limit comparability with other cohorts. Notably, the MMSE was originally developed as a screening tool for dementia and is relatively insensitive to deficits in executive functioning, attention, processing speed, and verbal memory, domains that are frequently affected in epilepsy.^48^ Consequently, participants with clinically meaningful cognitive difficulties may have scored within the normal range on the MMSE. Conversely, our use of the traditional cutoff score of 24 to define cognitive impairment may have overestimated impairment in this population, given its relatively low educational attainment. Prior studies, primarily using the Montreal Cognitive Assessment (MoCA), have suggested that lower cutoff scores may be more appropriate in rural populations with lower levels of education, as both the MoCA and MMSE were validated in highly educated populations.^49–51^ Unfortunately, comparable evidence for education-adjusted MMSE cutoffs is limited; therefore, we used the most widely studied conventional threshold. These considerations highlight the challenges of interpreting MMSE-based estimates of cognitive impairment in populations with substantial variation in educational attainment.

Another important limitation was the substantial missingness across several variables, which limited the ability to evaluate potentially important contributors to cognitive impairment, including psychiatric comorbidity and alcohol use. While mood and other psychiatric disorders have been consistently associated with subjective and objective cognitive difficulties among PWE, for example,^52–54^ we were unable to fully evaluate this variable due to high numbers of missing observations.

These limitations, however, must be considered in the context of the unique value of this clinical data. People with epilepsy in Latin America, and particularly those living in community settings, remain substantially understudied in research on cognitive outcomes. Despite incomplete ascertainment of some clinical characteristics, this large, community-based cohort provides important insight into cognitive function among people with epilepsy in a population that is rarely represented in the existing literature. As a population□based study, capturing both treated and untreated individuals with epilepsy in a *T. solium*□endemic region, selection bias, common in hospital□based studies, is minimized. The study also allows for a thorough clinical characterization of participants’ clinical history with variables including seizure type, treatment, and comorbidities, enabling examination of epilepsy-related drivers of cognitive impairment.

In conclusion, cognitive impairment was common among PWE in northern Peru and was associated with older age and lower educational attainment. This population represents a unique and vulnerable group in whom the cognitive burden of epilepsy may be compounded by limited access to neurologic, psychiatric, and cognitive care. The large size and community-based nature of this cohort provide an important perspective on cognitive functioning among PWE outside highly specialized clinical settings and may better reflect the population-level burden of cognitive impairment in regions with limited epilepsy care resources. These findings highlight the importance of recognizing and addressing cognitive health as part of comprehensive epilepsy care, particularly in resource-limited settings. Future studies using more detailed neuropsychological assessments and longitudinal designs are needed to better characterize the trajectory and determinants of cognitive impairment in these populations and to identify opportunities for intervention.

## Data Availability

All data produced in the present study are available upon reasonable request to the authors

