## Supplemental Table S1 for "Cognitive Impairment Among People with Epilepsy in Peru"

**Supplemental Table S1 –** Mini-Mental Status Exam (MMSE) Domain test components and scores

| **Domain** | **MMSE Component(s)** | **Score range** | **Mean, raw score (SD)** | **Mean, % of maximum (95% CI)** |
| --- | --- | --- | --- | --- |
| **Orientation** | *Temporal and spatial orientation* | *0-10* | 9.1 (1.6) | 91.7 (90.6-92.8) |
| **Registration** | *Immediate recall* | *0-3* | 2.9 (0.4) | 97.9 (97.1-98.8) |
| **Attention** | *Serial 7’s* | *0-5* | 4.0 (1.6) | 80.2 (77.9-82.6) |
| **Memory** | *Delayed recall* | *0-3* | 2.3 (1.0) | 77.3 (74.9-79.7) |
| **Language and praxis** | *Naming (2), repetition (1), three-step command (3), reading (1), writing (1), figure copying (1)* | *0-9* | 8.0 (1.6) | 89.1 (87.9-90.3) |
| **Total** | *All the above* | *0-30* | 26.3 (4.2) | 87.8 (86.8-88.8) |

**Abbreviations:** SD = standard deviation; CI = confidence interval
